# Trans-ancestry meta-analysis of genome wide association studies of inhibitory control

**DOI:** 10.1101/2022.10.13.22281074

**Authors:** Aurina Arnatkeviciute, Mathieu Lemire, Claire Morrison, Michael Mooney, Peter Ryabinin, Nicole Roslin, Molly Nikolas, James Coxon, Jeggan Tiego, Ziarih Hawi, Alex Fornito, Walter Henrik, Jean-Luc Martinot, Marie-Laure Paillère Martinot, Eric Artiges, Hugh Garavan, Joel Nigg, Naomi Friedman, Christie Burton, Russell Schachar, Jennifer Crosbie, Mark A. Bellgrove

## Abstract

Deficits in effective executive function, including inhibitory control are associated with risk for a number of psychiatric disorders and significantly impact everyday functioning. These complex traits have been proposed to serve as endophenotypes, however their genetic architecture is not yet well understood. To identify the common genetic variation associated with inhibitory control in the general population we performed the first trans-ancestry genome wide association study (GWAS) combining data across 8 sites and four ancestries (N=14,877) using behavioural traits derived from the stop-signal task, namely – go reaction time (GoRT), go reaction time variability (GoRT SD) and stop signal reaction time (SSRT). Although we did not identify genome wide significant associations for any of the three traits, GoRT SD and SSRT demonstrated significant and similar SNP heritability of 8.2%, indicative of an influence of genetic factors. Power analyses demonstrated that the number of common causal variants contributing to the heritability of these phenotypes is relatively high and larger sample sizes are necessary to robustly identify associations. The polygenic risk for ADHD was significantly associated with GoRT SD further supporting its suggested utility as an endophenotype for ADHD. Together these findings provide the first evidence indicating the influence of common genetic variation in the genetic architecture of inhibitory control quantified using objective behavioural traits derived from the stop-signal task.

## INTRODUCTION

Executive functions (EF) are essential in our everyday lives and critical for goal-directed behaviour. We need to adjust our actions based on changes in the environment, direct attention towards particular tasks, monitor performance, and inhibit irrelevant or automatic impulses. Broadly, these executive functions can be conceptualised as falling into three main categories – cognitive flexibility, working memory, and inhibitory control (1). Whereas EFs are linked to a range of positive outcomes such as educational attainment (2), quality of life (3,4) and general health-related behaviours (5), impairments in these cognitive processes are associated with risk for a number of psychiatric disorders including attention deficit hyperactivity disorder (ADHD) (6–8), autism spectrum disorder (9) as well as obsessive-compulsive disorder (OCD) (10–12) and schizophrenia (13,14).

Inhibitory control presents a particular facet of executive functioning that is directed at inhibiting inappropriate or irrelevant responses involving a set of distinct cognitive processes such as the ability to selectively control attention and behaviour as well as override the innate predisposition for a prompted action. Inhibitory control can be assessed in a laboratory setting using the stop signal paradigm (15,16), in which participants typically perform a “go” task but on a minority of the trials are presented with a stop signal that requires them to withhold an already initiated response to a go-signal. The performance in a stop-signal task is therefore modelled as a race between the initiated ‘go process’ that is triggered by a frequently presented go-stimulus and a ‘stop process’ which is triggered by the stop-signal, such that the response is inhibited if the stop process finishes before the go process (17). As a result, the performance on the stop signal task is characterised by three main measures: mean go reaction time (Go RT) reflecting the overall processing speed for go-stimuli, go reaction time variability (Go RT SD) corresponding to the efficiency with which top-down regulation of attention can be exerted over behaviour (18), and the stop signal reaction time (SSRT) which quantifies the efficiency of response inhibition, with longer SSRTs indicative of poorer response inhibition (15).

Deficits in inhibitory control and associated cognitive measures are at the core of ADHD symptoms such as impulsivity and inattention (19,20). Moreover, executive functions in general, and the measures of inhibitory control in particular, serve as the main candidate endophenotypes for ADHD (21–23). Just as ADHD has a strong genetic component (24–27), convergent evidence to date suggests that inhibitory control is also under substantial genetic influence with moderate heritability estimates ranging from h^2^=0.2-0.6 identified across a range of inhibitory control measures including the stroop task (28,29), stop signal task (28,30,31), go/no-go task (32), prohibition task (33), as well as the antisaccade task (28,34). Moreover, a latent variable derived from a combination of inhibitory control measures was almost entirely genetic in origin (28). Bivariate heritability analyses also indicate shared genetic influences between ADHD traits and the primary index of the efficiency of response inhibition derived from the stop-signal task, SSRT, suggesting the potential for common genetic contributions between the two phenotypes (30). Supplementing these behavioural findings, inhibition-related event components derived from electroencephalography (EEG) also demonstrate moderate heritability h^2^=0.5-0.6, further supporting the role of genetic influences in inhibitory control (35).

Quantifying the overall extent of genetic influences through heritability analyses provides the grounds for further investigations with the main goal of identifying the specific genes associated with inhibitory control that could help to determine contributing neurobiological mechanisms for these processes and associated disorders. Determining such genes so far has been a challenge with suggestive associations identified mainly through candidate gene studies linking the measure of response inhibition to genetic variations involving a number of genes such as the adrenergic receptor genes *ADRA2A* (36) and *ADRA2B* (37), norepinephrine transporter gene *SLC6A2* (38,39), dopamine transporter gene *DAT1* (40,41), dopamine receptor gene *DRD2* (42), serotonin type 2A receptor gene *HTR2A* (43), and neuronal tryptophan hydroxylase-2 gene *TPH2* (44). Candidate gene studies, however, have been extensively criticized due to high false-positive rates (45) and poor reproducibility (45,46). Indeed, a later study failed to identify any conclusive associations for any of seven *a priori* single nucleotide polymorphisms (SNPs) previously associated with stop signal task performance (47). Therefore, more systematic and powerful approaches are required to establish robust associations.

In contrast to candidate gene studies where genetic variants are selected *a priori*, genome-wide association studies (GWAS) provide a systematic approach to identifying genetic associations in a data-driven way, as well as allowing quantification of the extent of genetic influences attributable to common genetic variation. Several GWASs to date have investigated different aspects of executive functioning including processing speed (48–50), and the latent measures of working memory and inhibitory control (49), however very few genome-wide significant associations have been identified. The largest and most recent GWAS of executive function investigated the common executive functioning factor score (cEF) derived from multiple tasks in the UK Biobank dataset and found 129 independent lead variants mainly associated with fast synaptic transmission (51). SNP-heritability studies indicate that common genetic variation explains a substantial fraction of variance in working memory 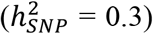 (49) and processing speed 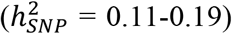 (48,49) suggesting that with enough power one can expect to identify more genome-wide significant associations that could inform the genetic mechanisms of different executive functions, including inhibitory control.

Here we performed the first trans-ancestry GWAS meta-analysis of inhibitory control in a general population sample of up to 14,877 participants, focusing on executive control measures derived from the stop-signal task. Go trial reaction time (GoRT) quantified processing speed, go reaction time variability (GoRT SD) quantified the efficiency of top-down regulation of attention, and stop signal reaction time (SSRT) served as a measure of response inhibition. Although we did not identify significant genome-wide hits for any of these phenotypes, the significant SNP heritability estimates for both response variability and response inhibition indicate that interindividual differences in both of these measures are influenced by genetic factors. Power analyses showed that in this study we had excellent power to detect at least one association at genome-wide significance if the number of common causal variants was ≤500. Our failure to identify genome-wide associations suggests that the actual number of contributing variants is significantly greater and larger sample sizes are necessary to identify robust associations. We also showed that the polygenic risk for ADHD was significantly associated with reaction time variability, further supporting the suggested utility of this measure as an endophenotype for ADHD.

## METHODS

### Participants

In this study we aggregated data across eight independent samples from the general population [SPIT1, SPIT2, Adolescent Brain Cognitive Development^SM^ Study (ABCD Study^**®**^), MELBOURNE, IMAGEN, COLORADO, Michigan-ADHD-1000, Oregon-ADHD-1000] and four ancestral groups [African (AFR), East Asian (EAS), European (EUR), South Asian (SAS)], totalling to 14,877 participants. Spit for science (SPIT1, SPIT2) is an ongoing study at The Hospital for Sick Children in Toronto (Canada) aiming to investigate the genetics of cognition, physical health and well-being in children aged 6-17 years (30,52). The ABCD Study is a publicly available longitudinal dataset from the USA containing participants aged 9 to 10 years at their baseline assessment, focusing on cognition, brain development, mental and physical health (53,54). The Melbourne sample (MELBOURNE) is derived from an ongoing study at Monash University in Melbourne, Australia that is designed to systematically assess neurocognition, psychopathological symptoms, genetics, as well as brain structure and function in a large sample of healthy young adults aged 18-50 years (mean 22.42, SD=4.89) (55). The IMAGEN sample was derived from the longitudinal IMAGEN dataset collected across eight centers in Europe combining brain imaging, genetics and psychiatry to understand brain development and behaviour in adolescents aged 14 years at baseline (39). The Colorado sample (COLORADO) includes same sex monozygotic (MZ) and dizygotic (DZ) twins recruited from the Colorado Longitudinal Twin Sample that was designed to investigate genetic and environmental influences on cognitive and emotional development (56,57). The Oregon-ADHD-1000 is a community-recruited (northwest Oregon, USA), longitudinal, case-control cohort of children (age 7-11 years at baseline) that is enriched for psychopathology (58–62). The Michigan-ADHD-1000 is a cohort of youth (age 6-21 years) with the same recruitment and assessment procedures as the Oregon-ADHD-1000 cohort, but recruited from a different demographic population (central Michigan, USA) (63,64). Only control subjects were selected for the analysis from both of the latter cohorts.

### Phenotypes

To investigate the genetics of executive function we selected three behavioural traits derived from the stop-signal task (SST) (65), namely, mean go reaction time (GoRT), go reaction time variability (GoRT SD) and stop signal reaction time (SSRT) representing overall processing speed, response variability, and response inhibition, respectively. All stop signal tasks consisted of two types of trials: “go” trials and “stop” trials. In a “go” trial participants are asked to respond to a stimulus as quickly and as accurately as possible by a button press corresponding to a particular stimulus. In “stop” trials participants are required to suppress their response to a go stimulus after the stop stimulus is presented therefore inhibiting an already initiated process. Stop signal tasks were administered independently between studies according to the site-specific study design and best practices (for the experimental procedures in each study, see Supplementary Text S1; for the description of SSRT integration method, see Supplementary Text S2).

### Genotyping and imputation

Samples were genotyped on a variety of arrays that are listed in Supplementary Table S1. For SPIT 1&2 studies, only participants for which all 4 grandparents shared the same ancestry (either EUR, EAS or SAS) were genotyped. For ABCD Study, we restricted analyses to non-Hispanic EUR, EAS, SAS and AFR ancestries. Recruitment for all other study cohorts was restricted to participants of EUR ancestry. Genotyping quality control (QC) was performed by different study centers according to their own best practice and pipelines (for genotyping and QC details for each site see Supplementary Text S3).

Imputation was performed separately for all studies and genotyping arrays, using ancestry-specific data from phase 3, version 5 of the 1000 Genomes project for reference. Data for SPIT 1&2 and ABCD Study[Go] were imputed using Beagle v4.1 (66). Data for MELBOURNE, IMAGEN and ABCD Study[SSRT] were imputed using minimac v4 on the Michigan imputation server (67). The COLORADO sample was imputed on the Michigan Imputation Server using minimac v4, Eagle v2.4 for phasing. Dosage data were used for these sites. For both the Oregon-ADHD-1000 and the Michigan-ADHD-1000, non-genotyped SNPs were imputed with the same procedure using IMPUTE2 (68); autosomal chromosomes were pre-processed and phased using SHAPEIT (69). Variant positions and alleles were checked against the reference panel and SNPs that were missing or mismatches were removed. Genotype probabilities for these two sites were converted to best-guess genotypes with genotype set to missing if the probability was <0.8.

### Association analysis

Association analyses were performed within each study and within each ancestral group, focusing on SNVs with MAF>1% and imputation quality r^2^>0.80. Most studies used allele dosage, while data in Oregon-ADHD-1000 and Michigan-ADHD-1000 samples were based on the best-guess genotype calls (i.e. from reading vcf files into plink). To account for the relatedness between some of the participants, we tested for association using the linear mixed models implemented in GEMMA v0.98.1 (70). All traits (mean GoRT, GoRT SD, SSRT) were analysed on the natural log scale. We used sex, age, age^2^ and age x sex as covariates, as well as the first 3 principal components constructed from the SNP data. An example from the SPIT1 study demonstrates that 3 principal components were sufficient to cluster regional ancestries within continental ancestries (see Supplementary Figure S1).

Within ancestral groups, the studies were meta-analysed using METAL release 2011-03-25 (71), with a focus on SNVs covering >70% of the samples, as was done elsewhere (24). Summary statistics from each site and ancestral group were meta-analysed using the methods described in (72) and originally implemented in MR-MEGA v0.1.5. Briefly, the method accounts for the possible heterogeneity of the effect sizes of a SNV in different ancestries by modelling in a regression framework the individual study effect sizes as a function of axes of genetic variation computed from multidimensional scaling. We used 3 axes of variation in addition to the regression intercept to model our 4 ancestral groups. For each SNP in study *s*, the observed effect size (*β*_*s*_) was estimated as:

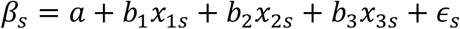

where *x*_1*s*_, *x*_2*s*_ and *x*_3*s*_ are the (pre-computed) values of study *s* in the 3 axes of variation (Supplementary Figure S2). Each study is weighted according to the inverse of the variance of its effect size. Significance is obtained from testing *a* = *b*_1_ = *b*_2_ = *b*_3_ = 0, in which case the observed effect sizes in each study are no different from random residuals (*∈*_*s*_). The original implementation of MR-MEGA can only analyse complete data, so we implemented our own regression in R to allow for missing results in some of the studies that arose due to frequency or imputation quality thresholds. We verified that results from our code and MR-MEGA agree for complete data. Axes of genetic variation were calculated using MR-MEGA from SNPs with complete data.

### SNP heritability, genetic correlations and polygenic scores

We assessed SNP heritability 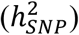 for each phenotype (mean GoRT, GoRT SD and SSRT) as well as the genetic correlation between each of those phenotypes using LD score regression as implemented in LDSC v1.0.0 (73). We restricted these analyses to SNVs with complete data to ensure that results were not affected by imbalances in power between studies. We used the LD scores as provided within LDSC v1.0.0 for EUR and EAS samples and performed our own calculations of scores for AFR and SAS samples using the same methods. We investigated trans-ancestry genetic correlations using POPCORN (installed from git commit #facdfbc)(74).

Polygenic scores (PGS) constructed from the ADHD PGC GWAS (24) were assessed for association in our samples with our traits using a pruning and thresholding approach as implemented in PLINK (75) and PRSice v1.25 (76), clumping SNPs for LD (using default *r*^*2*^<0.1 in 250 kb windows). PGS were evaluated at the p-value thresholds 0.001, 0.05, 0.10, 0.20, 0.30, 0.40, 0.50. We restricted these analyses to SNVs with imputation quality r^2^> 0.8. PGS effect sizes between studies were meta-analysed using fixed-effect, inverse variance methods. To account for testing multiple correlated PGS derived from the p-value inclusion thresholds, we calculated an effective number of independent PGS from the data and applied a Bonferroni correction with respect to that number (for a description, see Supplementary Text S4). We chose this approach of correcting for multiple testing because constraints on sharing individual level data precluded the use of permutation procedures.

## RESULTS

The total sample for each respective GWAS consisted of 14,844 subjects for GoRT SD, 14,877 for mean GoRT and 14,114 for SSRT (descriptive characteristics for each study are shown in Table 1). Samples from the different study centers and ancestries were generally comparable in terms of age and sex, with a few exceptions. ABCD Study had slightly younger participants with an age range that was narrower compared to other studies, whereas MELBOURNE and COLORADO studies consisted of young adult participants.

**Table 1.** Descriptive statistics for each ancestry and study sample. Sample sizes for each of the analysed phenotypes (GoRT SD, mean GoRT, and SSRT). Also shown are statistics for the covariates age and sex, the latter being expressed as a percentage of females. AFR – African ancestry; EAS – East Asian ancestry; EUR – European ancestry; SAS – South Asian ancestry.

| Ancestry | Study | N | N | N | Mean age | Females |
| --- | --- | --- | --- | --- | --- | --- |
|  |  | GoRT SD | Mean GoRT | SSRT | (SD) | % |
| AFR | ABCD Study | 781 | 781 | 706 | 9.95 (0.61) | 52 |
| EAS | ABCD Study | 97 | 97 | 89 | 9.91 (0.63) | 55 |
|  | SPIT1 | 847 | 847 | 847 | 11.51 (3.00) | 52 |
|  | SPIT2 | 294 | 294 | 294 | 10.26 (3.16) | 55 |
|  | **TOTAL** | **1238** | **1238** | **1230** |  |  |
| EUR | ABCD Study | 3844 | 3844 | 3577 | 9.95 (0.62) | 48 |
|  | SPIT1 | 4943 | 4943 | 4943 | 11.03 (2.75) | 48 |
|  | SPIT2 | 727 | 727 | 727 | 10.29 (3.03) | 49 |
|  | MELBOURNE | 942 | 942 | 668 | 22.42 (4.89) | 57 |
|  | IMAGEN | 1123 | 1123 | 1074 | 13.7 (3.39) | 48 |
|  | COLORADO | 524 | 524 | 524 | 22.59 (1.11) | 53 |
|  | OHSU | 159 | 159 | 155 | 9.43 (1.52) | 44 |
|  | MSU | 97 | 130 | 47 | 13.05 (3.21) | 48 |
|  | **TOTAL** | **12359** | **12392** | **11715** |  |  |
| SAS | ABCD Study | 32 | 32 | 29 | 10.05 (0.72) | 38 |
|  | SPIT1 | 250 | 250 | 250 | 11.52 (3.07) | 50 |
|  | SPIT2 | 184 | 184 | 184 | 10.51 (3.39) | 49 |
|  | **TOTAL** | **466** | **466** | **463** |  |  |
| **GRAND TOTAL** |  | **14844** | **14877** | **14114** |  |  |

### Association analyses

First, we performed trans-ancestry GWASs for each phenotype and found that no variant reached genome-wide significance (p<5×10^−8^) for any of the studied traits (Figure 1, Supplementary Figures S3-5 represent ancestry-specific analyses). Based on the investigation of LD score regression intercepts in the largest sample (EUR) we found that the potential biases caused by insufficiently controlled fine-scaled ancestry or cryptic relatedness were not significant for neither GoRT SD nor SSRT, indicating that association tests were not inflated (or deflated) (Table 2). Considering this result, the significant intercept deviation from 1 observed in the case of mean GoRT can be treated as spurious. Another possibility is that the genetic architecture of mean GoRT comprises predominantly rare causal variants, which are known to produce higher intercepts and negative slopes (73). In some cases, other ancestries also demonstrated intercepts exceeding 1 (depending on the trait), likely owing to admixture in these populations or small sample sizes (Table 2). Similar results were obtained using 10 PCs as covariates (not shown).

**Figure 1.**
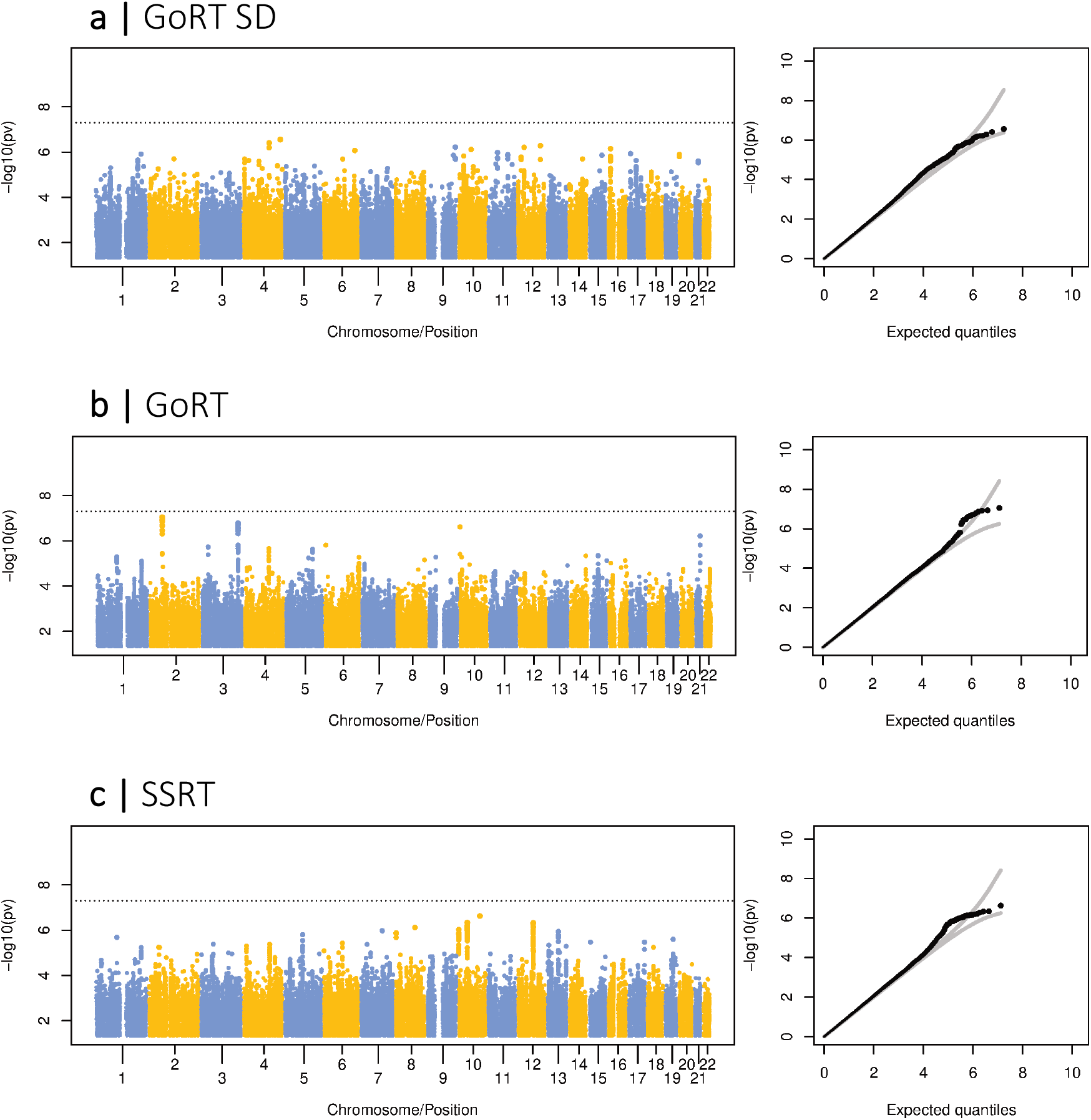
Trans-ancestry GWAS. Manhattan plots and corresponding quantile-quantile (QQ) plots for GoRT SD (a); mean GoRT (b); SSRT (c). Dashed lines on the Manhattan plots indicate p<5×10^−8^ threshold. Gray lines on the QQ plots represent 95% confidence bands.

**Table 2.** Heritability estimates for each phenotype and ancestry group. LD score regression estimates, standard error (SE) and significance (P-value) of the proportion of trait variance (*h*^2^) and regression intercept explained by common variants, stratified by ancestry. When the regression intercept is constrained, it is constrained to 1. Tests are one-sided for *h*^2^ (>0) and two-sided for the intercept (≠1). Heritability estimates presented in bold are considered statistically significant. AFR – African ancestry; EAS – East Asian ancestry; EUR – European ancestry; SAS – South Asian ancestry.

| Trait | Ancestry | h2 (constrained int.) |  |  | h2 (unconstrained int.) |  |  | Intercept |  |  |
| --- | --- | --- | --- | --- | --- | --- | --- | --- | --- | --- |
|  |  | Estimate | SE | P-value | Estimate | SE | P-value | Estimate | SE | P-value |
| **GoRT SD** | AFR | 0.157 | 0.150 | 0.147 | -0.230 | 0.214 | 0.858 | 1.018 | 0.007 | 0.015 |
|  | EAS | 0.369 | 0.258 | 0.076 | 0.243 | 0.402 | 0.273 | 1.006 | 0.008 | 0.675 |
|  | EUR | **0.082** | **0.029** | **0.002** | 0.065 | 0.050 | 0.096 | 1.005 | 0.012 | 0.675 |
| | SAS | 5.556 | 0.792 | $1 \times 10^{-12}$ | 0.221 | 1.093 | 0.420 | 1.047 | 0.008 | $2.7 \times 10^{-9}$ |
| **GoRT Mean** | AFR | 0.137 | 0.137 | 0.160 | -0.304 | 0.176 | 0.959 | 1.021 | 0.007 | 0.006 |
|  | EAS | 0.616 | 0.285 | 0.015 | -0.259 | 0.445 | 0.719 | 1.026 | 0.010 | 0.011 |
|  | EUR | 0.040 | 0.028 | 0.072 | -0.089 | 0.046 | 0.974 | 1.038 | 0.012 | 0.001 |
|  | SAS | 1.381 | 0.765 | 0.035 | 0.928 | 1.212 | 0.222 | 1.004 | 0.009 | 0.637 |
| **SSRT** | AFR | 0.008 | 0.716 | 0.496 | -0.258 | 0.940 | 0.608 | 1.002 | 0.006 | 0.713 |
|  | EAS | 0.574 | 0.266 | 0.016 | 0.599 | 0.396 | 0.065 | 0.999 | 0.009 | 0.940 |
|  | EUR | **0.081** | **0.031** | **0.004** | 0.008 | 0.053 | 0.440 | 1.020 | 0.013 | 0.125 |
| | SAS | 5.522 | 0.855 | $5.3 \times 10^{-11}$ | -1.003 | 1.114 | 0.816 | 1.059 | 0.008 | $4.3 \times 10^{-13}$ |

### Heritability and association with ADHD polygenic scores

Next, we evaluated the combined effect of common genetic variation for each phenotype by calculating SNP heritability 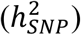 focusing on the largest available sample (EUR) (Table 2). Both GoRT SD and SSRT showed significant and similar SNP heritabilities of ∼8.2% (p=0.002 and p=0.004, respectively, when the intercept was constrained to reduce the variability). LD score regression intercept significantly departing from 1 would indicate a non-negligible impact of confounding factors such as cryptic relatedness and population stratification (73). In both cases, the intercept was not significantly different from 1, motivating the constraint. When the LD score intercept was free to vary, the point estimate for the GoRT SD was reasonably robust, albeit not significant 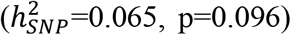, whereas for SSRT the effect of the constraint was critical (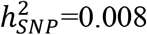 with unconstrained intercept, p=0.44). For completeness, Table 2 shows heritability for other ancestries, but owing to the relatively small sample sizes these should be viewed as non-informative and potentially misleading (77). We also investigated trans-ancestry genetic correlations between phenotypes in the two largest ancestral groups (EUR and EAS), however, due to small the sample sizes, the standard error of the trans-ancestry genetic correlation estimate (a parameter bounded by 1) was above 10, making inference and interpretation uninformative.

Deficits in executive function, including inhibitory control and response variability are associated with risk for ADHD (78,79). To evaluate the relationships between the genetic risk for ADHD and executive function we constructed polygenic scores for ADHD based on the PGC summary statistics (24) focusing the analyses on samples of EUR ancestry. The associations between ADHD PRS and each of the behavioural measures were performed in each study center separately and the effect sizes of the PGS (standardized to have unit variance) on the traits were meta-analysed. We found that ADHD PGS were significantly associated with GoRT SD, but did not show any associations with GoRT or SSRT (Figure 2, see Supplementary Table S2 for more detailed results). The largest and most significant effect of the PGS on GoRT SD (β=0.0079, se=0.0021; p=0.000123) was observed using a clumped set of SNPs retaining variants with p<0.5 based on the PGC ADHD GWAS, where larger PGS (representing the increased risk of ADHD) were associated with larger variability of the Go trial responses. This result was mostly driven by ABCD Study cohort (p=0.000126) and showed considerable (p=0.051) heterogeneity between studies. We calculated that the seven correlated PGS that were tested corresponded to an effective number of independent variables equal to ∼3 (*N*_*e*_=3.00 in ABCD Study; *N*_*e*_=2.96 in SPIT1), meaning that the above significance level for the PGS association with GoRT SD (p=0.0003) passes a Šidák correction for multiple testing even after accounting for the 3 traits that were tested [9 tests in total; p_sidak_=1-(1-0.0003)^9^=0.0026].

**Figure 2.**
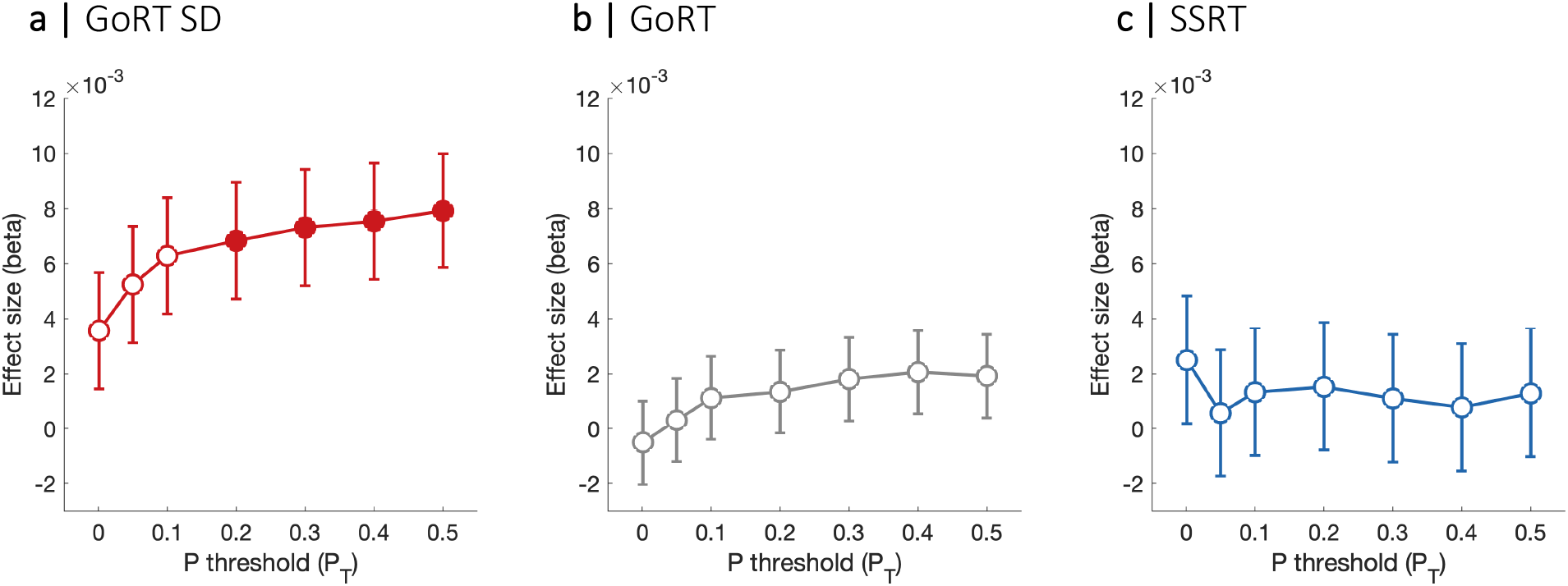
ADHD polygenic risk score associations. Associations between PRS for ADHD and GoRT SD (a), GoRT (b), and SSRT (c) based on the meta-analysis of EUR samples. Each subplot represents the estimated effect sizes (beta) and standard error (se) across a range of p-value thresholds (P_T_). Filled circles indicate association p-values that pass Šidák correction for multiple testing for the 3 traits (p<0.0026).

### Power analyses

In order to assess the power for detecting at least one association with a common (MAF>1% in EUR as baseline) causal variant (in this context, CV - the variant that is responsible for the association signal at a particular locus) at genome-wide significance, we performed a simulation study. Leveraging the significant and robust heritability for GoRT SD, we aimed to simulate a varying number of CVs, together explaining 8.2% of the variance of a simulated, normal trait. CVs were randomly selected among those with MAF>1% in the EUR population of the 1000 Genomes project and were assigned effect sizes drawn from a normal distribution and neutral selection. From a larger set of pre-simulated whole genomes, we randomly selected genotype data for 12,359 EUR, 1238 EAS, 466 SAS and 781 AFR samples, constructed the polygenic score from the causal ones and generated a trait by adding an environmental variance appropriately scaled (see Supplementary Text S5). For the effect sizes, we simulated two scenarios: one where the effect sizes are the same in all ancestries, and one where the effect sizes are uncorrelated between ancestries. CVs were taken to be the same, for parsimony. Details of the simulation designs are provided in the Supplementary Text S5.

We show that the simulated whole genomes are: i) indistinguishable from unrelated samples (Figure 3a), ii) that they closely preserve the LD structure of the original 1000 Genomes samples they are derived from (Figure 3b), and that iii) the simulated trait has the desired heritability, on average (Figure 3c). Under a model where the effect sizes are uncorrelated between ancestries, the trans-ancestry meta-analysis approach leads to a slightly reduced power compared to an analysis based only on samples from EUR ancestry, whereas comparable power is estimated for the model of correlated effect sizes (Table 3). These results are driven by the fact that the majority of samples in our study were derived from the EUR ancestry and are not necessarily the case for more balanced sample sizes. The loss of power in the trans-ancestry model in our case arises due to the estimation of three additional parameters (one per additional ancestry) (72) that due to relatively small sample sizes of the non-EUR ancestries are estimated with higher variability.

**Figure 3.**
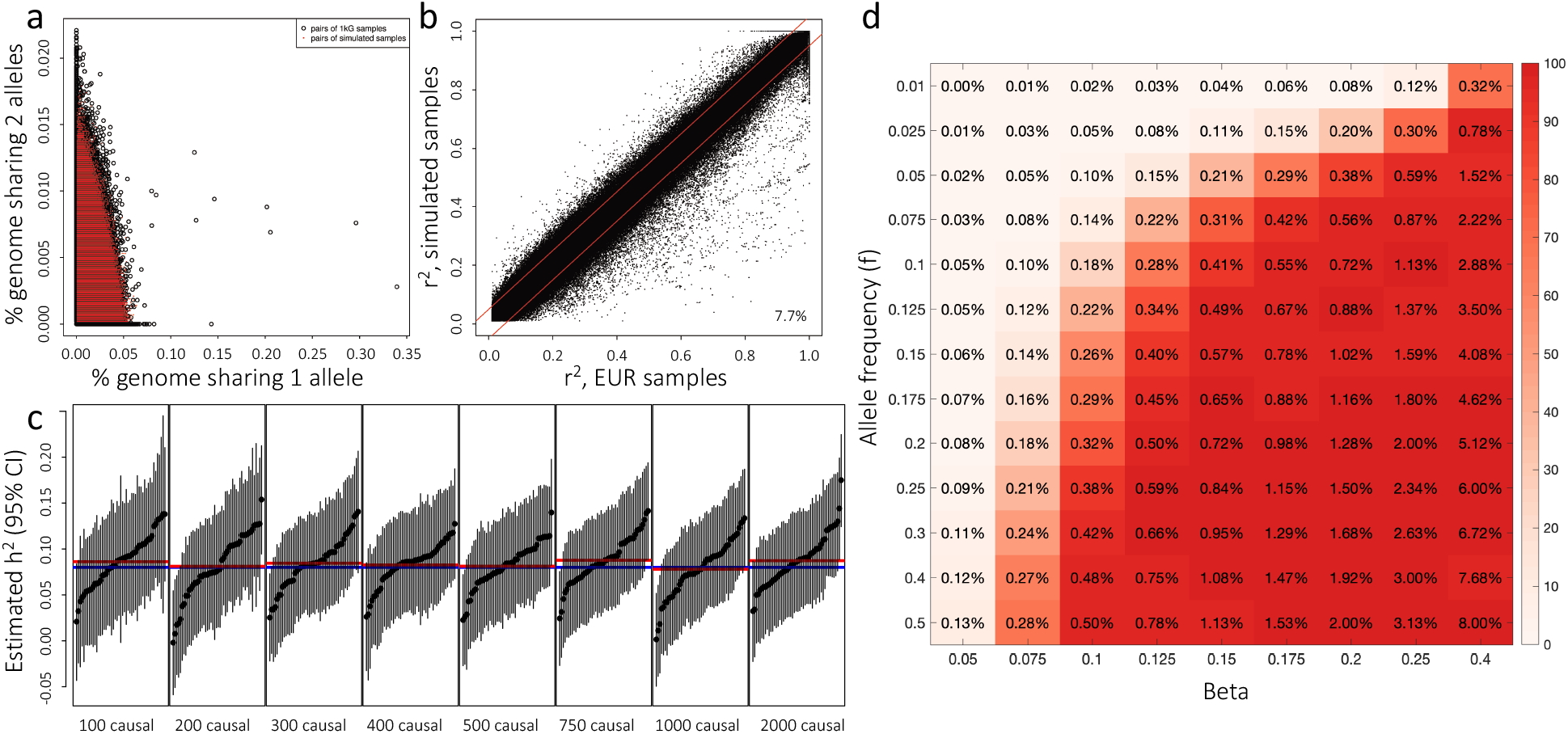
Validation of simulated replicates and the power to detect association for a single SNV. (a) Percentage of genome shared identical by descent between pairs of 503 EUR samples from the 1000 Genomes project (1kG; black circles) and between pairs of 10000 simulated samples derived from them (red dots). (b) Linkage disequilibrium (r^2^) between pairs of SNPs calculated in 503 EUR samples from 1kG (x-axis) compared to (size-matched) 503 simulated samples (y-axis). Red bands indicate differences of +/- 0.05; 7.7% of SNP pairs fall outside the bands. (c) Estimated LDSC heritability calculated from 12,359 simulated samples of EUR ancestry, for a trait simulated to have 8% heritability (blue horizontal line). Number of simulated causal variants are indicated on the horizontal axis. Red lines represent mean estimates, calculated from 100 simulated replicates. Vertical lines represent 95% confidence intervals for the heritability estimates (black points). (d) The power to detect association for a single SNV. The colours in the matrix represent the power (R2) to detect an association at genome-wide significance between a SNV and a unit-variance trait for varying allele frequency and effect size (beta: increase in trait value per minor allele). Values in each cell correspond to the percentage of trait variance explained by that SNV. R2 is calculated to be 2*Beta^2*f*(1-f).

**Table 3.** Simulation-based power calculations. Power to detect at least one association and the median number of discoveries at genome-wide significance p<5×10^−8^, as a function of the number (N) of causal variants (CVs). Each estimate is based on 200 simulated replicates. Between ancestries, effect sizes of the CVs were either correlated or uncorrelated. The simulated trait has LD-score heritability 8%.

| N | Correlated effect sizes |  | Uncorrelated effect sizes |  |
| --- | --- | --- | --- | --- |
|  | EUR only | Trans-ancestry | EUR only | Trans-ancestry |
| 100 | 100% (9) | 100% (10) | 100% (9.5) | 100% (9) |
| 200 | 100% (7) | 100% (7) | 100% (7) | 100% (6) |
| 300 | 100% (5) | 100% (5) | 100% (5) | 99% (3) |
| 400 | 96% (3) | 99% (3) | 96% (3) | 91% (2) |
| 500 | 94% (2) | 91% (2) | 91% (2) | 74% (1) |
| 750 | 69% (1) | 67% (1) | 66% (1) | 43% (0) |
| 1000 | 42% (0) | 41% (0) | 46% (0) | 24% (0) |
| 2000 | 14% (0) | 11% (0) | 13% (0) | 9% (0) |

Our results indicate that if the total number of common CVs explaining a LD score regression-derived 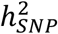 of 8.2% was ∼500 or less, then the power of our sample to detect at least one association at genome-wide significance level was excellent and generally above 80%, irrespective of the model or the method (Table 3). As a result, our failure to detect any association indicates that the number of CVs explaining 8.2% of the variance is likely to be more than ∼750-1000. When heritability is fixed, as the number of CVs increases, the proportion of trait variance explained by each variant decreases, resulting in decreasing power to detect any association. In our case, the power to detect an association with a particular SNV at genome-wide significance was adequate (>80%) as long as that SNV explained approximately >0.35% of the trait variance, which can be achieved for various combinations of MAF and effect sizes (Figure 3d). The fact that we did not detect any association, therefore, indicates that if a common causal SNVs was catalogued by the 1000 Genomes project, or unmeasured but in high LD with one, then this causal SNV is unlikely to explain more than ∼0.3% of a trait variance.

## DISCUSSION

Most quantifiable behavioral traits are termed complex due to the fact that they do not follow the Mendelian inheritance patterns; instead, they are influenced by a large number of genetic factors including multiple risk alleles, each of small effect size (80,81). Understanding the genetics of inhibitory control is critical for uncovering the genetic architecture of psychiatric and neurodevelopmental disorders such as ADHD that are characterized by significant impairments in a range of executive functions and inhibitory control in particular. Here we performed the first trans-ancestry GWAS using objectively-defined task-based measures of inhibitory control to investigate its genetic architecture. Although we did not identify any genome-wide significant variants, interindividual differences in measures of response inhibition (SSRT) and top-down regulation of attention (GoRT SD) were influenced by genetic factors. Critically, power analyses demonstrated that the lack of significant GWAS associations is due to the number of common causal variants contributing to the heritability of these phenotypes being relatively high and thus larger sample sizes are necessary to robustly identify associations. Linking inhibitory control to the genetics of ADHD we also identified a significant association between ADHD PGRS and reaction time variability, supporting its utility as an endophenotype for ADHD.

Considerable evidence from twin studies indicate moderate heritability for a range of inhibitory control measures (28–34), suggesting that in some tasks more than half of the variance in individual task performance can be explained by genetic factors. These relatively high values are in contrast to more modest heritability estimates accounting for the additive influence of common genetic variation in EFs based on GWASs that commonly do not exceed 30% (48–51). The discrepancy between twin and DNA-based measures is likely to be related to the effects of rarer genetic variants that are not assessed in GWAS, together with the nonadditive genetic effects (82), whereas another hypothesis suggests that the current estimates of twin-based heritability might be significantly inflated by genetic interactions (83). Here, for the first time we estimated a significant SNP-heritability for the measures of inhibitory control (GoRT SD and SSRT, 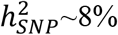), exceeding previous evaluations in a smaller sample of 4611 adolescents that failed to find common genetic contributions to stop signal task-based measures (49). Whereas our study similarly contained a large proportion of kids and adolescents (∼90%), the overall sample composition with regards to age could also impact heritability estimates as other cognitive domains tend to demonstrate increased influence of genetic factors later in life compared to childhood (84,85). Importantly, based on our simulations, we interpret the estimate of 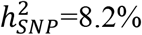 as the proportion of variance explained by common (>1%) SNVs catalogued by the 1000 Genomes project (or in high LD with these SNVs), but acknowledge that had we used a denser SNV imputation panel, the SNP heritability might have been higher (86). Whereas at the time the present project was initiated, the only available ancestry-diverse reference panel was from the 1000 Genomes project, the use of more recent ancestrally-diverse TOPMED reference panel (87) is encouraged for future research. Overall, our estimates were in line with the prior evidence of heritability of executive function (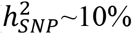 in largest samples) (48–51) indicating that the extent of common genetic influences on inhibitory control are comparable to more general factors of EF. Measures of executive function and inhibitory control in particular have been proposed as endophenotypes for ADHD (21–23). Our findings indicating the significant heritability and identifying the association between ADHD PRS and reaction time variability in a large sample through meta-analysis further support this idea. Whereas the initial search for endophenotypes was based on the assumption that these quantifiable traits should have less complex genetic architectures that are more closely related to gene function, (48–51), here we demonstrate the inherent complexity of genetic factors contributing to inhibitory control. Through power analyses we investigated the potential reasons why no genome-wide significant associations were identified, despite observing significant heritability of ∼8%. Our findings suggest that the number of common genetic variants explaining the identified heritability is likely to be relatively large exceeding 750-1000, each contributing not more than ∼0.3% of the variance. These estimates further support the contention that complex genetic architectures underly behavioural measures of response inhibition and top-down regulation of attention represented by SSRT and GoRT SD, respectively.

Currently the protocols for large-scale studies containing genomic data, such as UK Biobank, do not include measures of inhibitory control mainly due to the time required for data collection. Therefore, in order to achieve adequate sample sizes for a GWAS, data need to be aggregated across multiple studies. Challenges arise due to differences in experimental paradigms of the stop signal task with varying number of trials, individual trial lengths, mode of the stop stimuli (visual *vs* auditory), approaches for defining stop signal delay, as well as the methods used for measure estimation. Although it is not possible to retrospectively modify the individual study designs, here we aimed to control the variability in measure estimation by adhering to the best practice protocol proposed by Verbruggen et al., (2019), including exclusion of subjects that violate the assumptions of the race model, maintaining stop accuracy between 25%-75%, and use of the integration method for SSRT calculation where possible. To minimise variation in the genomic data all study sites used the same reference panel for imputation and imputation quality filter (r^2^>0.8). Nevertheless, despite implementing these measures in an attempt to standardise resulting calculations, some variation across study sites remained.

Historically most genomic research focused on genetically homogeneous cohorts from European populations limiting the generalisability of the identified findings and in some cases leading to biased inferences (88,89). Genomic data across different ancestral groups is however valuable and becoming increasingly available and will serve to increase the total sample sizes and representativeness of genetic studies. Nevertheless, integrating these data does pose some technical challenges as not all SNPs are polymorphic across different populations, some disease-associated SNPs have vastly different allele frequencies or show marked variability in linkage disequilibrium patterns with the causal variant between populations (90,91). Moreover, causal variants might interact with environmental risk factors that differ between ancestral populations additionally generating heterogeneity in the estimated effects. As a result, adjusting for population stratification opposes the goal of maximising the study power as traditional fixed and random effects approaches tend to under-estimate the effects sizes or over-estimate the standard errors reducing the overall confidence in the identified associations (92,93). Here we demonstrate the first attempt to incorporate data across different ancestries in the meta-analysis of inhibitory control using a method that derives the axes of genetic variation between populations based on genome-wide metrics of diversity via multi-dimensional scaling resulting in increased power over standard approaches while maintaining false positive error rates (72). Novel approaches for incorporating data from different ancestries are being continuously developed (72,94–96) providing opportunities for future large-scale trans-ancestry studies to uncover the genetic architecture of complex traits in a generalisable way.

In summary, in this first trans-ancestry GWAS of inhibitory control we demonstrated that task-derived measures of response inhibition and top-down regulation of attention are influenced by common genetic factors. Importantly, the number of contributing common genetic variants is likely to be relatively large suggesting that larger studies will be required to identify robust genome-wide associations. Our results also support the conceptualisation of reaction time variability as an endophenotype for ADHD providing grounds for targeting top-down regulation of attention through interventions.

## Data Availability

GWAS summary statistics data produced are available online at https://tinyurl.com/3w67mfyh.

## Code and data availability

Genotyping data processing code for SPIT1, SPIT2 and ABCD Study data is provided at http://bitbucket.org/mathieu-lemire/sk-scripts-qc-genotypingarrays. The custom MR-MEGA code implementation is provided at https://bitbucket.org/mathieu-lemire/sk_my_mrmega. Genotype simulation code is provided at https://bitbucket.org/mathieu-lemire/sk_recomb/src/master/; GWAS summary statistics for the trans-ancestry meta-analysis and EUR ancestry meta-analysis are provided at https://tinyurl.com/3w67mfyh;

## Acknowledgements

MAB was supported by a National Health and Medical Research Council of Australia Senior Research Fellowship. AA was funded by a grant from the Australian Research Council (ARC) under its Linkage Project scheme (LP160101592). A part of the data used in the preparation of this article were obtained from the Adolescent Brain Cognitive Development^SM^ (ABCD) Study (https://abcdstudy.org), held in the NIMH Data Archive (NDA). This is a multisite, longitudinal study designed to recruit more than 10,000 children age 9-10 and follow them over 10 years into early adulthood. The ABCD Study® is supported by the National Institutes of Health and additional federal partners under award numbers U01DA041048, U01DA050989, U01DA051016, U01DA041022, U01DA051018, U01DA051037, U01DA050987, U01DA041174, U01DA041106, U01DA041117, U01DA041028, U01DA041134, U01DA050988, U01DA051039, U01DA041156, U01DA041025, U01DA041120, U01DA051038, U01DA041148, U01DA041093, U01DA041089, U24DA041123, U24DA041147. A full list of supporters is available at https://abcdstudy.org/federal-partners.html. A listing of participating sites and a complete listing of the study investigators can be found at https://abcdstudy.org/consortium_members/. ABCD consortium investigators designed and implemented the study and/or provided data but did not necessarily participate in the analysis or writing of this report. This manuscript reflects the views of the authors and may not reflect the opinions or views of the NIH or ABCD consortium investigators. The ABCD data repository grows and changes over time. The ABCD genetic data and ABCD behavioural/cognitive data for GoRT and GoRT SD used in this report came from ABCD release 2.0; https://nda.nih.gov/study.html?id=634. The ABCD behavioural/cognitive data for SSRT used in this report came from ABCD release 3.0; https://nda.nih.gov/study.html?id=901.

