## Supplementary text for "Trans-ancestry meta-analysis of genome wide association studies of inhibitory control"

Aurina Arnatkeviciute*[1], Mathieu Lemire*[2], Claire Morrison[3,4], Michael Mooney[5,6], Peter Ryabinin[6], Nicole Roslin[2], Molly Nikolas[7], James Coxon[1], Jeggan Tiego[1], Ziarih Hawi[1], Alex Fornito[1], Walter Henrik[8], Jean-Luc Martinot[9], Marie-Laure Paillère Martinot[9,10], Eric Artiges[9,11], Hugh Garavan[12], Joel Nigg[13], Naomi Friedman[3], Christie Burton[2], Russell Schachar[2], Jennifer Crosbie*[2], Mark A. Bellgrove*[1]

*These authors contributed equally

[1]The Turner Institute for Brain and Mental Health, School of Psychological Sciences, Monash University, Australia

[2]Department of Psychiatry, The Hospital for Sick Children, Toronto, Canada

[3]Department of Psychology and Neuroscience, University of Colorado–Boulder, Boulder, CO, USA

[4]Institute for Behavioural Genetics, University of Colorado Boulder, Boulder, CO, USA

[5]Department of Medical Informatics & Clinical Epidemiology, Oregon Health & Science University, Portland, Oregon, USA

[6]Knight Cancer Institute, Oregon Health & Science University, Portland, Oregon, USA

[7]Department of Psychological and Brain Sciences, University of Iowa, Iowa City, IA, 52242, USA

[8]Department of Psychiatry and Psychotherapy, Charité-Universitätsmedizin Berlin, corporate member of Freie Universität Berlin and Humboldt-Universität zu Berlin, Germany

[9]Institut National de la Santé et de la Recherche Médicale, INSERM U1299 “Developmental trajectories & psychiatry” Université Paris-Saclay, Ecole Normale supérieure Paris-Saclay, CNRS, Centre Borelli, Gif-sur-Yvette, France

[10]AP-HP. Sorbonne Université, Department of Child and Adolescent Psychiatry, Pitié-Salpêtrière Hospital, Paris, France

[11]Etablissement Public de Santé (EPS) Barthélemy Durand, 91700 Sainte-Geneviève-des-Bois, France

[12]Departments of Psychiatry and Psychology, University of Vermont, 05405 Burlington, Vermont, USA

[13]Division of Psychology, Department of Psychiatry, Oregon Health & Science University, Oregon, Portland, USA

#### Supplementary Text S1

##### Phenotypes

*SPIT 1&2*

The full description of the task used in SPIT 1&2 samples can be found elsewhere (1). Briefly, the task consisted of a practice block (24 trials; 18 go trials; six stop trials) and four experimental blocks of 24 trials for a total of 72 go trials and 24 stop trials. Go trials consisted of the presentation of one of the two letters (an X or an O) on each trial where participants responded by pressing one key of a hand-held game pad for an X and the other for an O. The stop trials involved an auditory signal (1000Hz tone presented through headphones at a comfortable listening level) which was presented at random on 25% of all trials instructing participants to withhold the response on that particular trial. Each trial began with a fixation stimulus which was presented for 500ms followed by the go-signal (X or O) which was presented for 1000ms. The total duration of the trial was limited to 3500ms allowing 3000ms for a go task response. The task paused briefly after each block so that the supervisor could check if there were any error messages indicating that the participant was not following task instructions. Stop signal delay (SSD) was initially set at 250ms and then dynamically adjusted in 50ms increments depending on performance: after a successful stop trial the SSD was increased by 50ms (to make the stopping more difficult in the next trial) and after unsuccessful stop trials – shortened by 50ms.

Mean GoRT and GoRT SD were calculated based on trials that did not involve a stop signal. The SSRT was estimated using the integration method for subjects that: i) meet the racing assumption [such that the mean reaction time for failed stop trials was shorter than the mean reaction for Go trials] and ii) successfully perform 25%-75% of all stop trials. Go-reaction times in which no stop signal was presented were rank ordered and the Go-reaction time that corresponded to the probability of inhibition was determined. For example, if a participant inhibited 60 % of their Go-responses, one finds the 60th slowest Go-reaction time. All slower Go-responses would have been stopped; all faster ones would have been executed. Interpolated SSRT is estimated by subtracting mean delay from the integrated Go-reaction time (2).

*ABCD Study*

The full description of the SST in the ABCD Study can be found in (3). Briefly, the task consisted of 360 trials across 2 runs. Within each run 150 Go trials and 30 Stop trials were presented. Each trial lasted for 1000ms: Go trials comprised a response terminated arrow (50% rightward facing) followed by a fixation cross of variable length for a total trial duration of 1000ms; Stop trials comprised the arrow (50% rightward facing) presented for the duration of the variable stop signal delay (SSD) followed by a 300ms Stop Signal, and then by a fixation cross for a total duration of 1000ms. SSD was varied based on individual performance in 50ms increments. Stop trials are separated by 1 to 20 Go trials (mean 4.91) using twelve optimized trial orders. For a more detailed description see (4).

Data from ABCD Study release 2.0 were downloaded from http://dx.doi.org/10.15154/1503209. For Go reaction time and its variability, we selected the provided variables tfmri_sst_r1_beh_crgo_mrt (mean GoRT), tfmri_sst_all_beh_crgo_stdrt (GoRT SD) and restricted analysis to samples with acceptable performance in the task (tfmri_sst_beh_performflag=1). Poor performance was defined based on several criteria including fewer than 150 Go trials, less than 60% correct Go trials, more than 30% of incorrect Go trials, more than 30% of late Go trials (across correct and incorrect trials), more than 30% of nonresponsive Go trials, fewer than 30 Stop trials, and Stop trial accuracy lower than 20% or greater than 80%. SSRT measures estimated using the integration method were only available in release 3.0 (variable tfmri_sst_all_beh_total_issrt, therefore, for consistency with other measures, we selected only the subjects that were also available in release 2.0. Additional filters released in version 3.0 including the violation of the racing model assumption (tfmri_sst_beh_violatorflag=0), task coding errors (tfmri_sst_beh_glitchflag=0), the rate of incorrect Stop trials (0.25<tfmri_sst_all_beh_incrs_rt<0.75) and the omission rate of Go trials (tfmri_sst_all_beh_nrgo_rt>0.3) were implemented for quality control. Subjects with SSRT shorter than 120ms were removed.

*MELBOURNE*

Stop signal task (5) was presented in 12 blocks each consisting of 72 trials (54 go and 18 stop). Each trial lasted 1000ms: Go trials comprised a response terminated arrow (50% rightward facing) followed by a fixation cross of variable length for a total trial duration of 1000ms; Stop trials comprised the arrow (50% rightward facing) presented for the duration of the variable SSD followed by a 200ms Stop Signal, and then by a fixation cross for a total duration of 1000ms. SSD was varied based on individual performance in 50ms increments. Subjects that violated the racing assumption (such that the mean reaction time for failed stop trials was longer than the mean reaction for GO trials) were excluded. Go trial omissions and Go trials with response time <150ms were excluded before calculating the mean Go reaction time and its variability. SSRT was calculated using the integration method for blocks with not less than 25% (5/18) and not more than 75% (13/18) of successful stop trials. SSRT was then calculated for each block separately (6) and the median of SSRT values across blocks was taken to give a single SSRT value for an individual. Participants that had valid data from less than 5 out of 12 blocks as well as with SSRT shorter than 120ms were excluded.

*IMAGEN*

Stop signal task (7) consisted of 480 trials (400 go and 80 stop). Go trials presented a stimulus (arrows pointing left or right) for a duration of 1000ms. Stop trials comprised the go stimulus presented for a duration of variable SSD followed by a stop-signal (an arrow pointing upwards). Stop trials were separated by 3 to 7 Go trials. Stopping difficulty was manipulated across trials by varying SSD based on individual performance between 0 to 900ms in 50ms increments (initial SSD = 250ms). SSRT was calculated using integration method. Subjects with more than 80 errors in Go trials and SSRT shorter than 120ms were excluded.

*COLORADO*

The preparation for the stop signal task began with a single block of all-go trials (50 trials preceded by 10 practice trials) followed by a task practice for 48 trails containing both go and stop trials. The task was presented in 3 blocks each consisting of 80 trials (60 go and 20 stop) (8). In go trials green arrows were presented pointing left or right; in Stop trials the green arrow turned red at the varying SSD. Stopping difficulty was manipulated across trials using a staircase algorithm by varying SSD based on individual performance in 50ms increments (initial SSD = 200ms). Subjects that violated the assumptions of the race model (such that the mean reaction time for failed stop trials was longer than the mean reaction for GO trials) were excluded. SSRT was calculated using the integration method for blocks with not less than 25% and not more than 75% of successful stop trials and the mean across block SSRT values was calculated.

*Oregon-ADHD-1000 and Michigan-ADHD-1000*

Children completed the stop task in a single session lasting about 15-20 minutes. They sat facing a computer screen accompanied by a trained examiner who provided verbal instructions from a practiced script. The examiners were blind to child diagnosis and study hypotheses. The computer screen displays an X or an O on a black and white 14" computer monitor in random sequence. Children first complete two sets of 32 practice trials during which they practice pressing the designated key on the keyboard for the X or the O as quickly as possible, using two fingers on their dominant hand. They then completed a second set of 32 practice trials during which they again pressed the appropriate key, but this time trying not to hit any key when hearing a stop tone. Children are instructed to respond as quickly as possible without making errors on the X/O discrimination, that they will not be able in every instance to stop, and that they should not wait for the beep. The experimenter remained with each child throughout the task. In the experimental procedure, a fixation point appeared at the center of the screen for 500ms at the beginning of each trial. It was replaced by an X or O for 1000ms. The child responds with a key press, and the screen then is blank for 1000ms. Four blocks of 64 trials were administered, with a rest in between each block, for a total of 256 trials. On 25% of the trials, the stop tone sounded for 100ms. Thus, a total of 64 trials (16 per block) included the stop signal tone. The timing of the stop tone was varied to implement the tracking procedure as follows. The SSD was initially set at 250ms. If the child failed to inhibit their response after the tone, on the subsequent trial the SSD was decreased by 50ms (making it easier to stop next time); if they inhibited successfully, SSD was increased by 50ms on the next trial (making it harder to stop next time). Probability of stopping successfully is thus maintained at approximately 50%.

Prior to creating the total score across blocks, the following validity criteria were applied to each block: a) stop accuracy between 30-70%, b) hit accuracy greater than 75%, and c) mean RT for the block between 100-1500ms (to avoid anticipations on current or next trial). Mean go response time (GoRT) was calculated by averaging the correct go-trial response times across all blocks. SSRT, the primary measure of response inhibition, was calculated for each valid block by the integration method following previously published guidelines (9); SSRTs less than 50ms were considered invalid. The practice trials and the first block of data were removed to exclude warm-up effects. Valid block SSRT scores were averaged to create the final outcome variable.

#### Supplementary Text S2

##### Stop signal reaction time calculation using integration method

The integration method (9,10) for estimating the stop signal reaction time is based on the "integration" of the reaction time distribution and identifying the point at which the area under the curve is equal to the probability of response in stop trials [p(respond|signal)]. This way the SSRT corresponds to the nth RT, where the n is the number of go RTs in the distribution multiplied by the p(respond|signal). For instance, if there are 300 go trials and the probability of response to a stop trial is 0.45 [p(respond|signal) = 0.45], then the nth RT is the 135$^{th}$ (300 x 0.45) fastest go RT. SSRT is then estimated as a difference between nth RT and mean stop signal delay (SSD – the time between the presentation of a go and stop stimuli). For the most accurate estimate, go trials should include all trials with a response while replacing go omissions (trials where a participant did not respond) with the maximum RT to compensate for the missing response.

#### Supplementary Text S3

##### Genotyping and genotyping QC

SPIT1, SPIT2 and ABCD Study data were processed using the same pipeline deposited at http://bitbucket.org/mathieu-lemire/sk-scripts-qc-genotypingarrays. In brief, samples from a given study and given genotyping array were first stratified based on their ancestry when needed (based on self-report and/or the first 3 principal components – in which case a sample was assigned the ancestry of the nearest sample from the 1000 Genomes project). QC procedures were applied to each stratum separately. Samples were excluded if their call rate was below 97%, a heterozygosity rate of 6 times the interquartile range from the closest quartile and/or their predicted and reported sex were mismatched. One copy of duplicated samples was retained based on highest call rate. Monozygotic twin pairs were split – only the member with the lowest alphanumerical ID was retained. Subjects genotyped on plate number 461 in the ABCD Study dataset were removed due to poor data quality as recommended in the ABCD Study documentation. SNPs were excluded if they had call rates below 97%, they deviated from the rules of Hardy-Weinberg equilibrium at an FDR <1% and/or were duplicates of other SNPs, based on position and alleles (only the SNP with the highest call rate was retained).

The quality control for the MELBOURNE and IMAGEN study centers involved initially excluding subjects with very low-genotyping score (>10% of missing data) and SNPs with genotyping call rate <90%. SNPs with a minor allele frequency (MAF) < 0.01 were also removed. Further, individuals with: i) disparities between the recorded and observed sex status; ii) with low genotyping score (>5% of missing data); iii) cryptic relatedness higher than 0.25 and iv) displaying outlying mean heterozygosity (greater than ±3 SDs from the sample mean) were excluded. To identify potential sources of population stratification multidimensional scaling (MDS) was performed using HapMap3 dataset. Consequently, subjects exceeding ±2 SD on the 1st or 2nd principal components were excluded. SNPs with low genotyping call rate <95%, MAF< 0.01 and significantly departing from Hardy-Weinberg (HW) equilibrium ($p< 10^{-7}$) were removed before imputation.

The COLORADO sample was genotyped on the Axiom Precision Medicine Research array 2.0 chip or the Affymetrix 6.0 chip. QC consisted of removing indels, rare variants (MAF<0.01), variants with a high missing call rate (GENO<0.02) and variants out of Hardy-Weinberg equilibrium ($p<10^{-4}$). Those with cryptic relatedness >0.2 were removed after computing a GRM.

The Oregon-ADHD-1000 and Michigan-ADHD-1000 cohorts were both genotyped using the Illumina PsychChip v1.1. For the Oregon cohort, all individuals had genotype call rates >98%. SNPs were excluded if they had a call rate <97%, Hardy-Weinberg equilibrium deviation ($p<10^{-6}$), ambiguous strand information, or differential call rates between genotyping batches or ethnic groups. SNPs located in regions of suggestive copy number anomalies as calculated by B-allele frequency were also removed. Additional SNPs determined by Illumina to perform poorly across populations were excluded. For the Michigan cohort, all individuals had a genotyping rate >97%. Genotyped SNPs were excluded from analysis if their call rate was < 97%, had Hardy-

Weinberg equilibrium p-value $<10^{-5}$, had a call rate difference between ADHD cases and controls >2%, had a batch effect FDR adjusted p-value < 0.05, or were discordant among duplicate samples.

#### Supplementary Text S4

##### Multiple testing correction for PGS estimation

An effective number of independent PGS ($N_e$) can be calculated as (11,12):

$$N_e = 1 + (N_t - 1)\left(1 - \frac{Var(\lambda)}{N_t}\right),$$

where $N_t$ is the number of p-value thresholds used (here, 7) and $\lambda$ are the eigenvalues derived from the $N_t$ PGS (centered and scaled to unit-variance). If all PGS were uncorrelated, then all eigenvalues would take the value 1 and have no variance, leading to $N_e$=$N_t$, whereas if all were perfectly correlated, the first eigenvalue would take the value $N_t$ while all others are 0, leading to $N_e$=1. Since the overall correlation between the PGS (more generally, variables) can be captured by the variance of the eigenvalues, the expression represents the proportional reduction in the number of PGS after accounting for their correlations (11,12).

#### Supplementary Text S5

##### Simulated whole genomes

The program consists of a collection of scripts that uses plink and UNIX tools to simulate whole genomes that are "descendants" of samples from the 1000 Genome project. An alternative and viable choice for the simulations presented here would have been to use HAPGEN2 (13), but we wanted the flexibility of designing a tool that we could easily expand to simulate sibs or other relatedness, or ad-mixed samples for other research projects. We started with the samples from the 1000 Genomes project (phase 3 version 5; downloaded from http://bochet.gcc.biostat.washington.edu/beagle/1000_Genomes_phase3_v5a/b37.vcf/) as the founders of our simulated samples. We extracted haplotypes from the phased diplotype data and stored them in plink format (these are thus stored as homozygous genotypes). Haplotypes were randomly paired, and recombination breakpoints were randomly chosen based on an interpolated, sex-averaged genetic map (http://compgen.rutgers.edu/rutgers_maps.shtml). The recombinant haplotypes could then serve to construct the next generation, even though we did not simulate generations *per se*: for illustration, we calculated that, in order to grow the 503 EUR from the 1000 Genomes into a population of 100,000 simulated samples, 15 generations would be sufficient (assuming a growth rate of 1.5% per year and 25 years per generation). The genetic length of chromosome 1 is about 280 cM, or an average of 2.8 recombination per meiosis (generation). Following one lineage over the course of 15 generation, this translates to 42 recombination events on chromosome 1. For simplicity, instead of simulating 15 generations of mating, we simulated 42 recombinations, randomly selecting haplotypes between each event (which increases diversity). For other chromosomes, the number of events was proportional to the genetic lengths. The number 42 of recombination events was fixed and not an average over a probabilistic model. The idea was not to mimic a proper genealogy and a probabilistic recombination model that is as close as possible to the underlying biology, but rather introduce enough recombination events in order to end up with sets of unrelated samples, preserving as much of the original linkage disequilibrium as possible. This has the advantage of being easy to implement in a script: we only used plink to split and recombine data on both sides of a breakpoint, as well as bcftools, vcftools and standard UNIX commands. Using a two-hundred node computer cluster, we simulated 100,000 whole genomes over the course of 3 days. Our scripts are available in a package named RECOMB available at https://bitbucket.org/mathieu-lemire/sk_recomb/src/master/.

**Simulated trait values**

We randomly selected N causal variants (CVs) among the ones having MAF>1% in the EUR samples from the 1000 Genomes project. Each were independently assigned an effect size drawn from a normal distribution with mean 0 and variance 1, denoted $\beta_i$ for the minor allele of the $i^{th}$ CV. The additive genetic score at these N variants of an individual having $g_i$ mean-centered minor alleles at $CV_i$ is:

$$A = \sum_i g_i \beta_i.$$

To end up with a trait T that has heritability $h^2$, A was complemented with a normally distributed environmental residual *E* that has mean 0 and variance equal to $Var(A)*(1-h^2)/h^2$:

$T = A + E.$

Trait values can then be standardized to have unit variance if desired. In our simulations, the heritability was fixed for EUR (our only ancestry with a significant heritability estimate). For other ancestries, the heritability was free to differ according to the allelic spectra of the causal variants. In order to do so, the variance of E for other ancestries was taken to be the value calculated in EUR. We surmise that the variance of E is driven mostly by the experimental conditions, which should not vary based on the ancestry of the participant performing the task.

Some (14–16) have used standardized effect sizes $\beta_i/\sqrt{2f_i(1-f_i)}$ in lieu of $\beta_i$ (where $f_i$ are allele frequencies); i.e.

$$A = \sum_i g_i\beta_i/\sqrt{2f_i(1-f_i)}.$$

This increases the range of effect sizes for lower frequency variants, motivated by selective pressures but also for optimality of the LD score regression framework (14). Since the $\beta_i$ are independent with mean 0 and variance 1 and independent of $g_i$, and the variance of $g_i$ is $2f_i(1-f_i)$, then under that model the variance of A is equal to N, irrespective of the allelic frequencies of the causal variants, and thus irrespective of the ancestries. This means that the heritability would be the same in all ancestries unless we arbitrarily and artificially set it to different values by varying Var(E). This motivated our choice for using an un-standardised (neutral selection) model for effect sizes. We surmise that our traits are not under strong selective pressures.

**GWAS with simulated traits and genomes**

The simulated trait and genomes were assessed for association as described in the main text. Since trait values were not simulated to be associated with covariates, covariates were not included in the analyses (alternatively, trait can be viewed as residuals after covariate adjustment). The simulated sample sizes were taken to be the ones from Table 1 (GoRT SD). CVs were assumed to be ungenotyped and were removed from the analyses. A CV was deemed to have been "discovered" if at least one SNV in LD with it ($r^2>0.5$ in at least one ancestry) reached genome-wide significance $p<5\text{x}10^{-8}$ (for computational speed we only analysed SNVs in LD with CVs, except for $h_{SNP}^2$ validation from LDSC where all HapMap3 SNPs were analysed). Power was defined to be the proportion of simulated replicates where at least one CV was discovered at genome-wide significance. The number of causal variants (N) varied from 100 to 2000; 200 simulated replicates were used for each N.

### SUPPLEMENTARY FIGURES

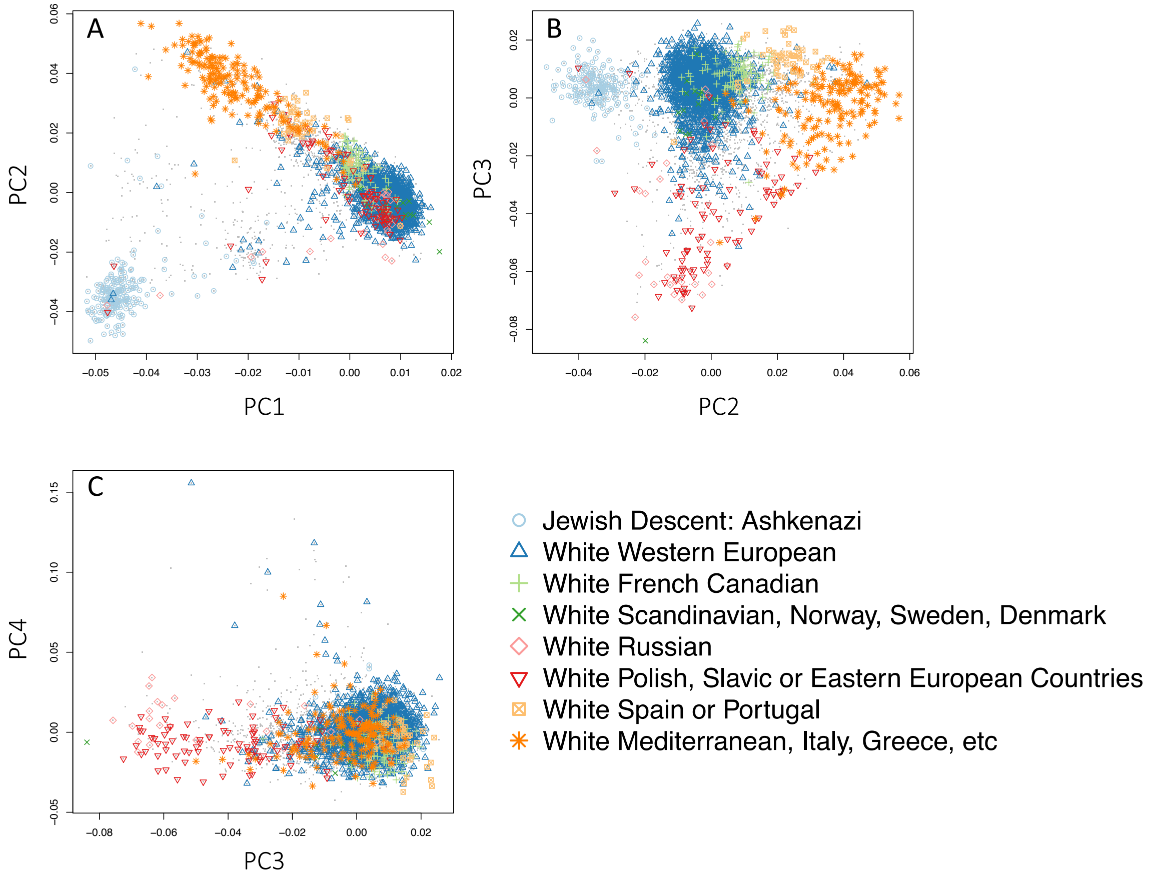


Figure S1. **Fine scale EUR ancestry in SPIT1 sample.** First four principal components representing the fine scale population structure: A) PC1 *vs* PC2; B) PC2 *vs* PC3; C) PC3 *vs* PC4. The different symbols indicate the common ancestry (self-reported) of the four grandparents of the participants. Dots without symbols represent participants whose four grandparents do not share the same fine-scale European ancestry or have missing data. No additional ancestry clusters can be derived from the fourth component and likely represent technical variations.

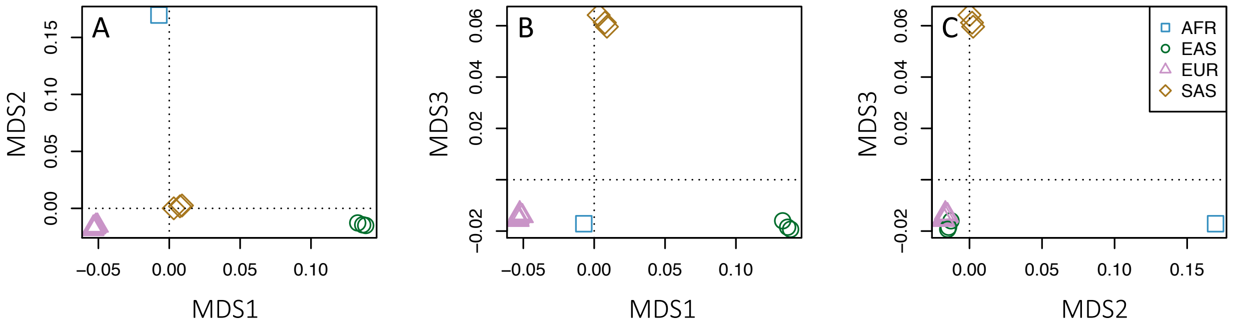


Figure S2. **Multidimensional scaling.** Three axes of genetic variations (MDS) used in the trans-ethnic regression framework, calculated with MR-MEGA from allele frequencies derived from summary statistics: A) MDS1 *vs* MDS2; B) MDS1 *vs* MDS3; C) MDS2 *vs* MDS3. Each point represents an individual study, color-coded based on participant ancestry.

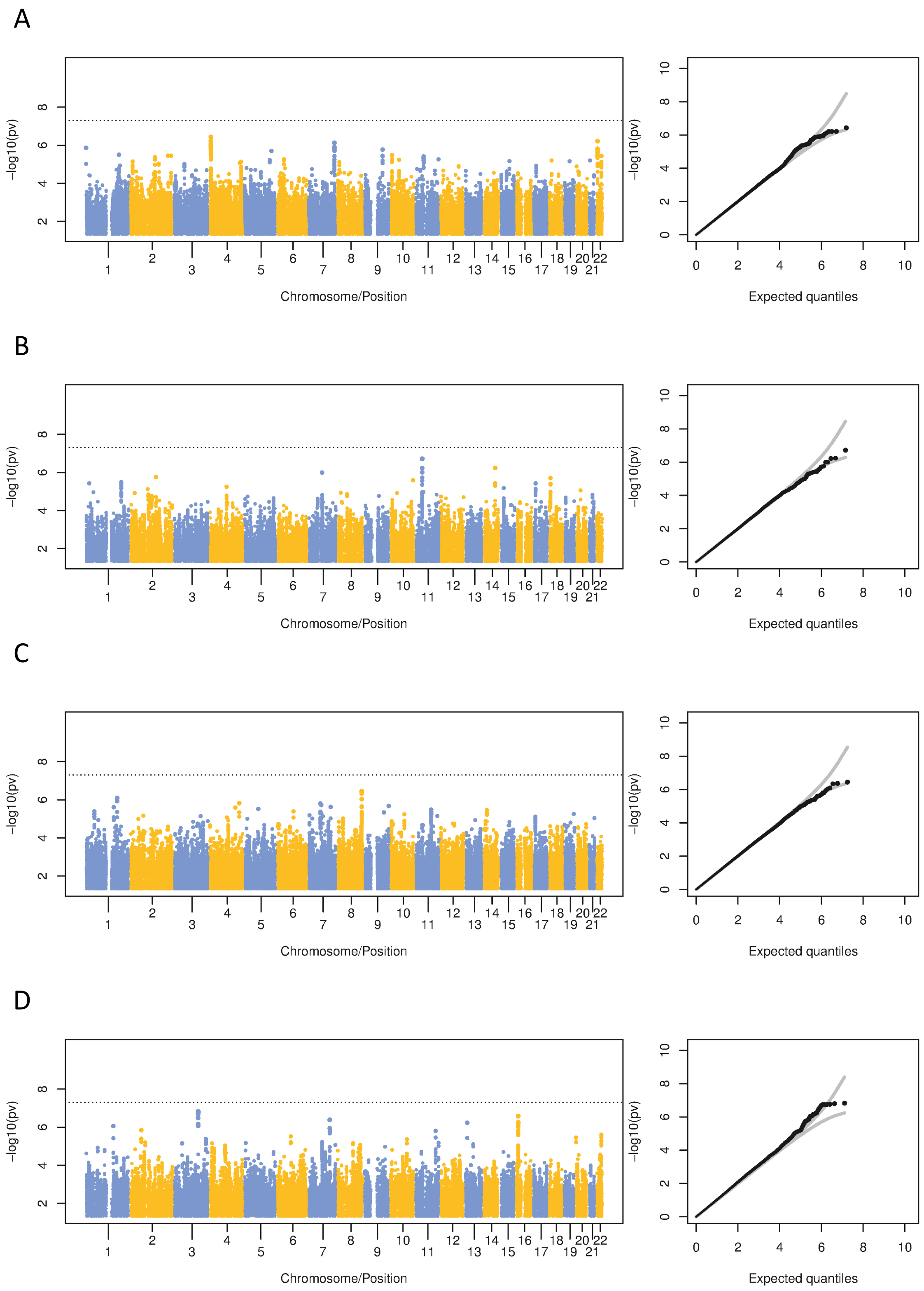


Figure S3. **GWAS for GoRT SD by ancestry.** Manhattan plots and corresponding qq-plot for (A) AFR; (B) EAS; (C) EUR; (D) SAS. Dashed lines on the Manhattan plots indicate $p<5x10^{-8}$ threshold. Gray lines on the QQ plots represent 95% confidence bands.

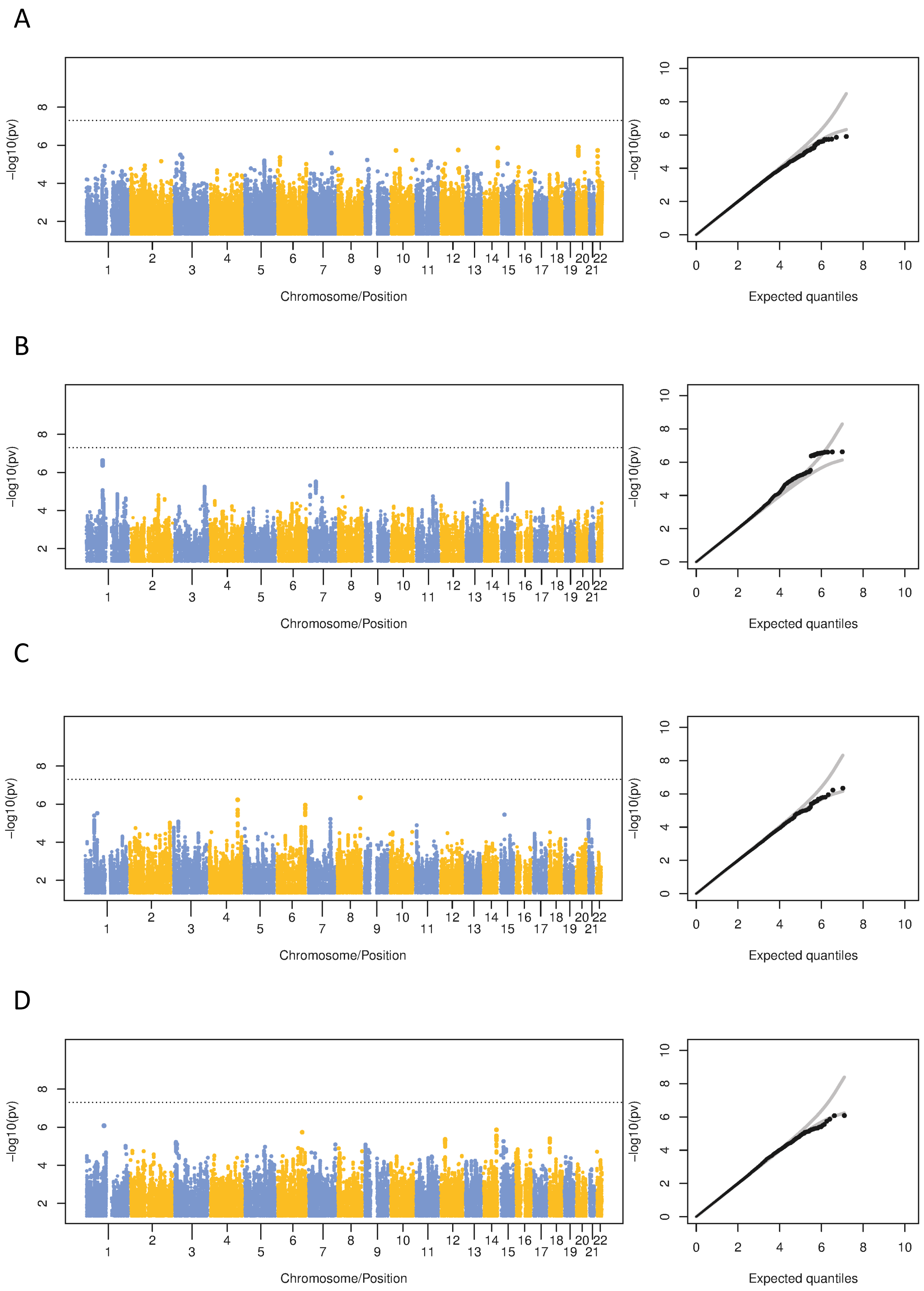


Figure S4. **GWAS for mean GoRT by ancestry.** Manhattan plots and corresponding qq-plot for (A) AFR; (B) EAS; (C) EUR; (D) SAS. Dashed lines on the Manhattan plots indicate $p<5x10^{-8}$ threshold. Gray lines on the QQ plots represent 95% confidence bands.

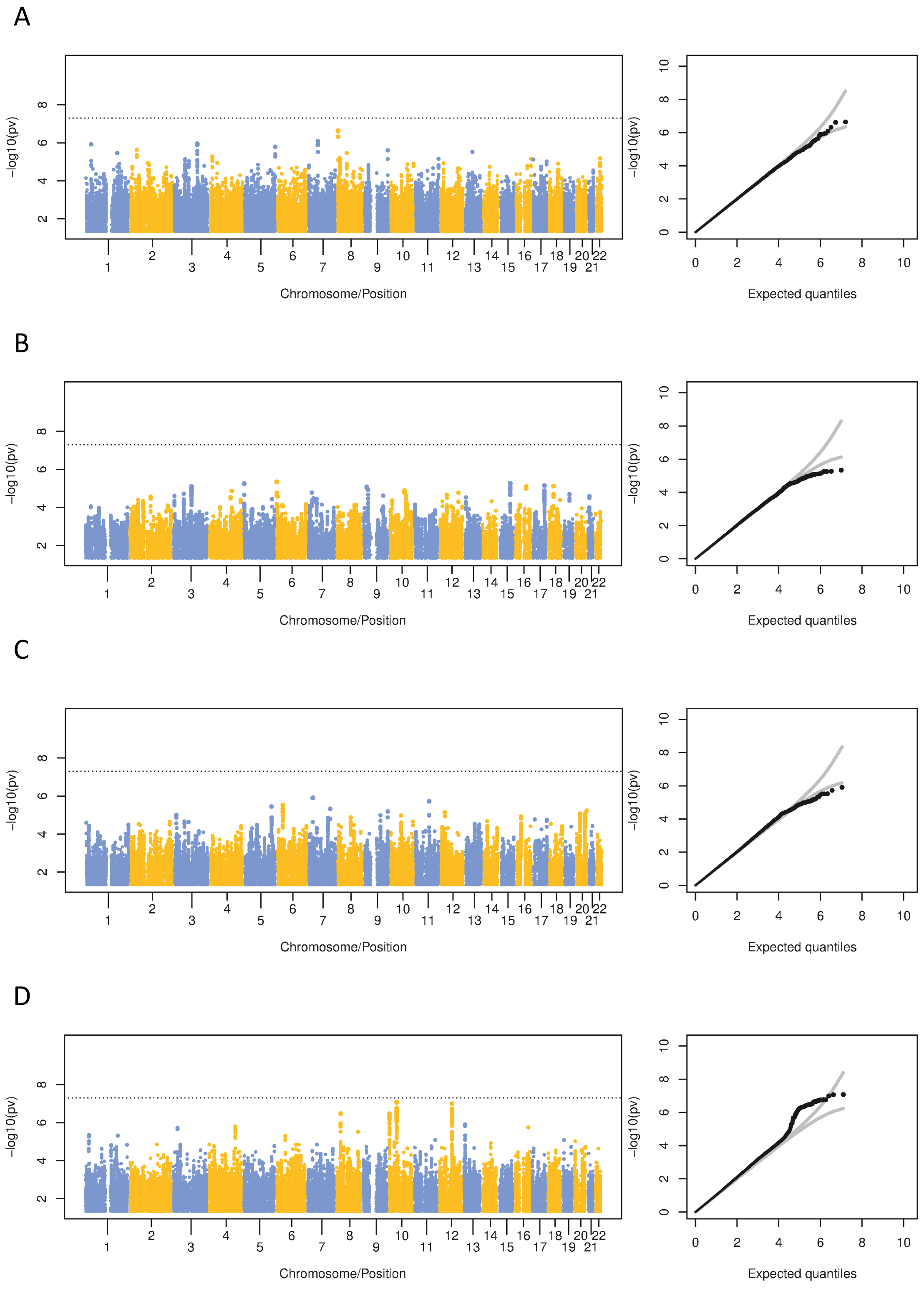


Figure S5. **GWAS for SSRT by ancestry.** Manhattan plots and corresponding qq-plot for (A) AFR; (B) EAS; (C) EUR; (D) SAS. Dashed lines on the Manhattan plots indicate p<5x10-8 threshold. Gray lines on the QQ plots represent 95% confidence bands.

#### SUPPLEMENTARY TABLES

Supplementary Table S1. Genotyping arrays used in each study.

| Study | Genotyping arrays |
|---|---|
| ABCD Study | Affymetrix NIDA SmokeScreen Array |
| SPIT1 | Illumina HumanCoreExome-12v1.0_B (EAS, EUR)<br>Illumina HumanOmni1-Quad V1.0_B (192 EUR)<br>Illumina GlobalDiversityArray-8v1-0_A1 (SAS) |
| SPIT2 | Illumina GlobalScreeningArray-24v3-0_A1 |
| MELBOURNE | Illumina Innium PsychArray-24v1.2 BeadChip |
| IMAGEN | Illumina Human610-Quad Beadchip and Illumina Human660-Quad Beadchip |
| COLORADO | Axiom Precision Medicine Research Array 2.0 |
| OHSU | Illumina PsychArray v1-1 |
| MSU | Illumina PsychArray v1-1 |
