## Supplementary table for "Trans-ancestry meta-analysis of genome wide association studies of inhibitory control"

| Trait | Study | P-value threshold | | | | | | | | | | | | | | | | | | | | |
|---|---|---|---|---|---|---|---|---|---|---|---|---|---|---|---|---|---|---|---|---|---|---|
| | | **0.001** | | | **0.05** | | | **0.1** | | | **0.2** | | | **0.3** | | | **0.4** | | | **0.5** | | |
| | | **beta (se)** | **R2** | **p** | **beta (se)** | **R2** | **p** | **beta (se)** | **R2** | **p** | **beta (se)** | **R2** | **p** | **beta (se)** | **R2** | **p** | **beta (se)** | **R2** | **p** | **beta (se)** | **R2** | **p** |
| **GoRT SD** | ABCD | 0.0018 (0.0027) | <0.001 | 0.5 | **0.0081 (0.0027)** | **0.0027** | **0.00217** | **0.0087 (0.0027)** | **0.0031** | **0.00102** | **0.0096 (0.0027)** | **0.0038** | **0.000299** | **0.0096 (0.0027)** | **0.0038** | **0.000276** | **0.0099 (0.0027)** | **0.004** | **0.0002** | **0.0102 (0.0026)** | **0.0042** | **0.000126** |
| | SPIT1 | 0.0028 (0.0042) | <0.001 | 0.511 | -0.0001 (0.0042) | <0.001 | 0.973 | 0.0024 (0.0042) | <0.001 | 0.572 | 0.0023 (0.0042) | <0.001 | 0.591 | 0.0039 (0.0042) | <0.001 | 0.363 | 0.0033 (0.0042) | <0.001 | 0.439 | 0.0033 (0.0042) | <0.001 | 0.433 |
| | SPIT2 | 0.0098 (0.0108) | <0.001 | 0.368 | -0.0118 (0.0108) | <0.001 | 0.276 | -0.0064 (0.0108) | <0.001 | 0.556 | -0.0019 (0.0108) | <0.001 | 0.859 | -0.0008 (0.0108) | <0.001 | 0.943 | 0.0009 (0.0108) | <0.001 | 0.937 | 0.0014 (0.0108) | <0.001 | 0.894 |
| | MEL | 0.0100 (0.0109) | <0.001 | 0.36 | -0.0123 (0.0110) | 0.0012 | 0.265 | -0.0124 (0.0110) | 0.0013 | 0.259 | -0.0129 (0.0110) | 0.0014 | 0.243 | -0.0115 (0.0110) | 0.0011 | 0.296 | -0.0098 (0.0110) | <0.001 | 0.374 | -0.0075 (0.0110) | <0.001 | 0.498 |
| | IMA | 0.0222 (0.00948) | 0.00485 | 0.0195 | 0.0292 (0.00947) | 0.0084 | 0.0021 | 0.0273 (0.00946) | 0.00738 | 0.00394 | 0.0257 (0.00948) | 0.00654 | 0.00668 | 0.0248 (0.00948) | 0.00606 | 0.00904 | 0.0268 (0.00948) | 0.0071 | 0.00471 | 0.0268 (0.00948) | 0.00707 | 0.0048 |
| | OHSU | -0.0095 (0.0228) | <0.001 | 0.676 | -0.0304 (0.0229) | 0.008 | 0.186 | -0.0319 (0.0229) | 0.0088 | 0.166 | -0.0407 (0.0232) | 0.014 | 0.081 | -0.0378 (0.0232) | 0.012 | 0.104 | -0.0382 (0.0231) | 0.012 | 0.1 | -0.0375 (0.0231) | 0.012 | 0.107 |
| | MSU | 0.0001 (0.0345) | <0.001 | 0.997 | 0.0076 (0.0340) | <0.001 | 0.823 | -0.0032 (0.0351) | <0.001 | 0.926 | 0.0005 (0.0353) | <0.001 | 0.989 | -0.0004 (0.0351) | <0.001 | 0.991 | -0.0063 (0.0348) | <0.001 | 0.857 | -0.0138 (0.0348) | 0.0012 | 0.692 |
| | | **beta (se)** | **p_het** | **p** | **beta (se)** | **p_het** | **p** | **beta (se)** | **p_het** | **p** | **beta (se)** | **p_het** | **p** | **beta (se)** | **p_het** | **p** | **beta (se)** | **p_het** | **p** | **beta (se)** | **p_het** | **p** |
| | META | 0.00357 (0.00211) | 0.5 | 0.0904 | **0.00524 (0.00211)** | **0.0109** | **0.013** | **0.00629 (0.00211)** | **0.0331** | **0.00285** | **0.00684 (0.00211)** | **0.0267** | **0.0012** | **0.00731 (0.00211)** | **0.0598** | **0.000531** | **0.00754 (0.00211)** | **0.0454** | **0.000352** | **0.00792 (0.00206)** | **0.0511** | **0.000123** |
| | | **0.001** | | | **0.05** | | | **0.1** | | | **0.2** | | | **0.3** | | | **0.4** | | | **0.5** | | |
| | | **beta (se)** | **R2** | **p** | **beta (se)** | **R2** | **p** | **beta (se)** | **R2** | **p** | **beta (se)** | **R2** | **p** | **beta (se)** | **R2** | **p** | **beta (se)** | **R2** | **p** | **beta (se)** | **R2** | **p** |
| **GoRT Mean** | ABCD | -0.0035 (0.0025) | <0.001 | 0.166 | -0.0020 (0.0025) | <0.001 | 0.416 | -0.0021 (0.0025) | <0.001 | 0.408 | -0.0025 (0.0025) | <0.001 | 0.308 | -0.0020 (0.0025) | <0.001 | 0.414 | -0.0020 (0.0025) | <0.001 | 0.426 | -0.0022 (0.0025) | <0.001 | 0.373 |
| | SPIT1 | 0.0002 (0.0024) | <0.001 | 0.935 | 0.0012 (0.0024) | <0.001 | 0.61 | 0.0033 (0.0024) | <0.001 | 0.171 | 0.0040 (0.0024) | <0.001 | 0.104 | 0.0046 (0.0024) | <0.001 | 0.058 | 0.0048 (0.0024) | <0.001 | 0.051 | 0.0047 (0.0024) | <0.001 | 0.056 |
| | SPIT2 | -0.0049 (0.0061) | <0.001 | 0.425 | -0.0011 (0.0061) | <0.001 | 0.862 | 0.0013 (0.0061) | <0.001 | 0.828 | 0.0040 (0.0061) | <0.001 | 0.51 | 0.0052 (0.0061) | <0.001 | 0.397 | 0.0051 (0.0061) | <0.001 | 0.398 | 0.0049 (0.0061) | <0.001 | 0.417 |
| | MEL | 0.0023 (0.0069) | <0.001 | 0.742 | -0.0114 (0.0070) | 0.0027 | 0.101 | -0.0101 (0.0070) | 0.0021 | 0.146 | -0.0076 (0.0070) | 0.0012 | 0.274 | -0.0068 (0.0070) | <0.001 | 0.331 | -0.0060 (0.0070) | <0.001 | 0.388 | -0.0052 (0.0070) | <0.001 | 0.457 |
| | IMA | 0.00920 (0.00468) | 0.00338 | 0.0494 | 0.01175 (0.00468) | 0.0055 | 0.0121 | 0.00954 (0.00467) | 0.00364 | 0.0414 | 0.00792 (0.00468) | 0.0025 | 0.091 | 0.00760 (0.00468) | 0.0023 | 0.1051 | 0.00841 (0.00468) | 0.00282 | 0.0728 | 0.00825 (0.00468) | 0.00271 | 0.0784 |
| | OHSU | -0.0118 (0.0170) | 0.0029 | 0.49 | -0.0186 (0.0171) | 0.0072 | 0.279 | -0.0152 (0.0172) | 0.0048 | 0.378 | -0.0267 (0.0174) | 0.014 | 0.127 | -0.0281 (0.0173) | 0.016 | 0.106 | -0.0256 (0.0173) | 0.013 | 0.14 | -0.0285 (0.0172) | 0.017 | 0.1 |
| | MSU | 0.0032 (0.0167) | <0.001 | 0.849 | 0.0108 (0.0168) | 0.0028 | 0.521 | 0.0110 (0.0172) | 0.0027 | 0.522 | 0.0188 (0.0172) | 0.008 | 0.275 | 0.0146 (0.0170) | 0.0049 | 0.39 | 0.0197 (0.0169) | 0.009 | 0.246 | 0.0186 (0.0169) | 0.008 | 0.273 |
| | | **beta (se)** | **p_het** | **p** | **beta (se)** | **p_het** | **p** | **beta (se)** | **p_het** | **p** | **beta (se)** | **p_het** | **p** | **beta (se)** | **p_het** | **p** | **beta (se)** | **p_het** | **p** | **beta (se)** | **p_het** | **p** |
| | META | 0.000502 (0.00152 | 0.321 | 0.741 | 0.000317 (0.00152) | 0.0759 | 0.835 | 0.00113 (0.00152) | 0.146 | 0.458 | 0.00135 (0.00152) | 0.088 | 0.375 | 0.00181 (0.00152) | 0.102 | 0.234 | 0.00207 (0.00152) | 0.0883 | 0.174 | 0.00193 (0.00152) | 0.0806 | 0.205 |
| | | **0.001** | | | **0.05** | | | **0.1** | | | **0.2** | | | **0.3** | | | **0.4** | | | **0.5** | | |
| | | **beta (se)** | **R2** | **p** | **beta (se)** | **R2** | **p** | **beta (se)** | **R2** | **p** | **beta (se)** | **R2** | **p** | **beta (se)** | **R2** | **p** | **beta (se)** | **R2** | **p** | **beta (se)** | **R2** | **p** |
| **SSRT** | ABCD | 0.00735 (0.00430) | <0.001 | 0.0872 | 0.00222 (0.00423) | <0.001 | 0.5998 | 0.00376 (0.00424) | <0.001 | 0.3744 | 0.00507 (0.00424) | <0.001 | 0.232 | 0.00394 (0.00424) | <0.001 | 0.353 | 0.00239 (0.00425) | <0.001 | 0.5732 | 0.00295 (0.00425) | <0.001 | 0.4878 |
| | SPIT1 | 0.0016 (0.0062) | <0.001 | 0.79 | -0.0000 (0.0062) | <0.001 | 0.999 | -0.0018 (0.0062) | <0.001 | 0.765 | -0.0030 (0.0062) | <0.001 | 0.623 | -0.0028 (0.0062) | <0.001 | 0.648 | -0.0030 (0.0062) | <0.001 | 0.627 | -0.0029 (0.0062) | <0.001 | 0.642 |
| | SPIT2 | 0.0085 (0.0150) | <0.001 | 0.573 | -0.0035 (0.0150) | <0.001 | 0.814 | 0.0014 (0.0150) | <0.001 | 0.927 | 0.0082 (0.0150) | <0.001 | 0.585 | 0.0073 (0.0150) | <0.001 | 0.629 | 0.0089 (0.0150) | <0.001 | 0.554 | 0.0123 (0.0150) | <0.001 | 0.412 |
| | MEL | 0.0049 (0.0039) | 0.0023 | 0.215 | 0.0024 (0.0039) | <0.001 | 0.536 | 0.0034 (0.0039) | 0.0011 | 0.386 | 0.0035 (0.0040) | 0.0011 | 0.38 | 0.0031 (0.0040) | <0.001 | 0.437 | 0.0034 (0.0040) | 0.0011 | 0.396 | 0.0044 (0.0040) | 0.0018 | 0.265 |
| | IMA | -0.00875 (0.00539) | 0.002448 | 0.105 | -0.00493 (0.00540) | <0.001 | 0.361 | -0.00465 (0.00539) | <0.001 | 0.388 | -0.00523 (0.00540) | <0.001 | 0.333 | -0.00505 (0.00540) | <0.001 | 0.35 | -0.00508 (0.00540) | <0.001 | 0.347 | -0.00552 (0.00540) | <0.001 | 0.307 |
| | OHSU | -0.0087 (0.0245) | <0.001 | 0.722 | 0.0126 (0.0249) | 0.0017 | 0.612 | 0.0097 (0.0249) | 0.001 | 0.696 | 0.0022 (0.0253) | <0.001 | 0.932 | 0.0035 (0.0252) | <0.001 | 0.89 | 0.0042 (0.0251) | <0.001 | 0.868 | 0.0015 (0.0251) | <0.001 | 0.954 |
| | MSU | -0.0535 (0.0586) | 0.018 | 0.367 | -0.0261 (0.0555) | 0.0049 | 0.641 | 0.0056 (0.0563) | <0.001 | 0.921 | 0.0063 (0.0568) | <0.001 | 0.913 | -0.0004 (0.0580) | <0.001 | 0.995 | 0.0180 (0.0572) | 0.0022 | 0.755 | 0.0302 (0.0573) | 0.0062 | 0.601 |
| | | **beta (se)** | **p_het** | **p** | **beta (se)** | **p_het** | **p** | **beta (se)** | **p_het** | **p** | **beta (se)** | **p_het** | **p** | **beta (se)** | **p_het** | **p** | **beta (se)** | **p_het** | **p** | **beta (se)** | **p_het** | **p** |
| | META | 0.00251 (0.00231) | 0.293 | 0.279 | 0.000579 (0.00230) | 0.924 | 0.802 | 0.00133 (0.00231) | 0.899 | 0.565 | 0.00154 (0.00233) | 0.777 | 0.507 | 0.00111 (0.00233) | 0.86 | 0.634 | 0.000782 (0.00233) | 0.866 | 0.737 | 0.00130 (0.00233) | 0.731 | 0.577 |

**Polygenic score analysis.** Polygenic scores (PGS) constructed from a GWAS for ADHD in EUR samples from sets of (clumped) SNPs reaching varying P-value threshols in the GWAS. Effect sizes (beta), standard errors (se) and significance (p) are indicated, along with the proportion of the trait variance explained by the PGS (R2); for the meta-analysis of the study centers, significance of tests of heterogeneity (p_het) are given in lieu of R2.
